# Statistical Issues and Lessons Learned from COVID-19 Clinical Trials with Lopinavir-Ritonavir and Remdesivir

**DOI:** 10.1101/2020.06.17.20133702

**Authors:** Guosheng Yin, Chenyang Zhang, Huaqing Jin

**Affiliations:** Department of Statistics and Actuarial Science, The University of Hong Kong, Hong Kong

**Keywords:** Coronavirus, COVID-19, Cure rate model, Sample size adjustment, Terminal event, Type I error rate, Restricted mean survival time

## Abstract

**Background:** Since the outbreak of the novel coronavirus disease 2019 (COVID-19) in December 2019, it has rapidly spread in more than 200 countries or territories with over 8 million confirmed cases and 440,000 deaths by June 17, 2020. Recently, three randomized clinical trials on COVID-19 treatments were completed, one for lopinavir-ritonavir and two for remdesivir. One trial reported that remdesivir was superior to placebo in shortening the time to recovery, while the other two showed no benefit of the treatment under investigation. However, several statistical issues in the original design and analysis of the three trials are identified, which might shed doubts on their findings and the conclusions should be evaluated with cautions.

**Objective:** From statistical perspectives, we identify several issues in the design and analysis of three COVID-19 trials and reanalyze the data from the cumulative incidence curves in the three trials using more appropriate statistical methods.

**Methods:** The lopinavir-ritonavir trial enrolled 39 additional patients due to insignificant results after the sample size reached the planned number, which led to inflation of the type I error rate. The remdesivir trial of Wang et al. failed to reach the planned sample size due to a lack of eligible patients, while the bootstrap method was used to predict the quantity of clinical interest conditionally and unconditionally if the trial had continued to reach the originally planned sample size. Moreover, we used a terminal (or cure) rate model and a model-free metric known as the restricted mean survival time or the restricted mean time to improvement (RMTI) in this context to analyze the reconstructed data due to the existence of death as competing risk and a terminal event. The remdesivir trial of Beigel et al. reported the median recovery time of the remdesivir and placebo groups and the rate ratio for recovery, while both quantities depend on a particular time point representing local information. We reanalyzed the data to report other percentiles of the time to recovery and adopted the bootstrap method and permutation test to construct the confidence intervals as well as the *P* values. The restricted mean time to recovery (RMTR) was also computed as a global and robust measure for efficacy.

**Results:** For the lopinavir-ritonavir trial, with the increase of sample size from 160 to 199, the type I error rate was inflated from 0.05 to 0.071. The difference of terminal rates was −8.74% (95% CI [-21.04, 3.55]; *P*=.16) and the hazards ratio (HR) adjusted for terminal rates was 1.05 (95% CI [0.78, 1.42]; *P*=.74), indicating no significant difference. The difference of RMTIs between the two groups evaluated at day 28 was −1.67 days (95% CI [-3.62, 0.28]; *P*=.09) in favor of lopinavir-ritonavir but not statistically significant. For the remdesivir trial of Wang et al., the difference of terminal rates was −0.89% (95% CI [-2.84, 1.06]; *P*=.19) and the HR adjusted for terminal rates was 0.92 (95% CI [0.63, 1.35]; *P*=.67). The difference of RMTIs at day 28 was −0.89 day (95% CI [-2.84, 1.06]; *P*=.37). The planned sample size was 453, yet only 236 patients were enrolled. The conditional prediction shows that the HR estimates would reach statistical significance if the target sample size had been maintained, and both conditional and unconditional prediction delivered significant HR results if the trial had continued to double the target sample size. For the remdesivir trial of Beigel et al., the difference of RMTRs between the remdesivir and placebo groups up to day 30 was −2.7 days (95% CI [-4.0, −1.2]; *P*<.001), confirming the superiority of remdesivir. The difference in recovery time at the 25th percentile (95% CI [-3, 0]; *P*=.65) was insignificant, while the differences manifested to be statistically significant at larger percentiles.

**Conclusions:** Based on the statistical issues and lessons learned from the recent three clinical trials on COVID-19 treatments, we suggest more appropriate approaches for the design and analysis for ongoing and future COVID-19 trials.

## Introduction

The novel coronavirus disease 2019 (COVID-19) has spread all over the world at an unprecedented rate since its outbreak in December 2019. More than 200 countries or territories have confirmed cases, and a total of over 8 million individuals have been infected, leading to more than 44,0000 deaths by June 17, 2020. The COVID-19 was declared as a public health emergency of international concern by the World Health Organization (WHO) on January 30, and as a pandemic on March 11, 2020.

As recommended by the WHO R&D Blueprint expert group, clinical improvements for COVID-19 patients can be classified as seven-category ordinal scales [1]:

1. Not hospitalized with resumption of normal activities;
2. Not hospitalized, but unable to resume normal activities;
3. Hospitalized, not requiring supplemental oxygen;
4. Hospitalized, requiring supplemental oxygen;
5. Hospitalized, requiring nasal high-flow oxygen therapy, noninvasive mechanical ventilation, or both;
6. Hospitalized, requiring ECMO, invasive mechanical ventilation, or both;
7. Death.

So far, there are only eight clinical trials for COVID-19 completed with results published. Among them, two trials were for hydroxychloroquine with relatively small sample sizes (30 patients for the trial of Chen et al. [2] and 36 patients for the trial of Gautret et al. [3]). Although the trial conducted by Gautret et al. [3] yielded a significant result, the sample size was too small to draw any convincing conclusion. The trial of Cai et al. [4] compared favipiravir and lopinavir-ritonavir with a total sample size of 80 patients, leading to a significant result with *P*=.004. Chen et al. [5] conducted a trial comparing favipiravir with arbidol, which had a total sample size of 240 patients and yielded an insignificant result. The trial of Grein et al. [6] was a single-arm trial for remdesivir and the estimated clinical improvement rate at day 18 was 0.68. We take the three randomized clinical trials conducted by Cao et al. [7] on lopinavir-ritonavir and by Wang et al. [8] and Beigel et al. [9] on remdesivir as examples to illustrate statistical issues and lessons learned from them as they have drawn great attention in the clinical community.

### Lopinavir-ritonavir trial

The trial LOTUS China (Lopinavir Trial for Suppression of SARS-Cov-2 in China) [7] was conducted at a record speed from January 18 to February 3, 2020 (the date of enrollment of the last patient), although patient recruitment up to a planned sample size is often the bottle neck of trial conduct. This was not the case with severe COVID-19 due to abundance of hospitalized patients during that period of time. In this trial, eligible patients were randomized at a 1:1 ratio to either the lopinavir-ritonavir treatment group (400 mg and 100 mg orally, twice daily) plus the standard care, or the standard care alone, for 14 days. No placebo was used for blinding because no placebo was prepared due to the urgency of the trial, so that both patients and investigators were aware of the identity of treatment each patient received. Following the WHO seven ordinal scales [1], the primary end point adopted by the trial [7] was the time to clinical improvement, which was defined as the time from randomization to an improvement of two points from the status at randomization (e.g., from point 6 to point 4 or from point 5 to point 3) or live discharge from the hospital, whichever came first. The sample size was increased from 160 to 199 since the result with the enrolled 160 patients did not reach statistical significance. As a final conclusion, Cao et al. [7] reported no benefit with the lopinavir-ritonavir treatment beyond the standard care with a hazard ratio (HR) of 1.24 and the associated 95% confidence interval (CI) [0.90,1.72].

### Remdesivir trial 1

Wang et al. [8] conducted a randomized, double-blind, placebo-controlled, multicentre trial with remdesivir at ten hospitals in Hubei, China. Overall, 236 patients were enrolled from February 6 to March 12, 2020, who were randomly assigned to the remdesivir group (200 mg on day 1 followed by 100 mg on days 2–10) and the placebo group at a 2:1 ratio. In the original design, it planned to recruit 453 patients with 302 to remdesivir and 151 to placebo, while no patients were enrolled after March 12 due to no more eligible patients available in the Hubei province. As a consequence, the statistical power of the study was reduced from 80% to 58%. The primary clinical end point was the time to improvement within 28 days. Clinical improvement was defined as two-point improvement from an adjusted six scales from the WHO seven-category ordinal scales. In conclusion, remdesivir did not show statistically significant clinical benefit compared with the placebo in terms of the HR of 1.23 (95% CI [0.87, 1.75]).

### Remdesivir trial 2

Beigel et al. [9] reported a randomized, double-blind, placebo-controlled trial of intravenous remdesivir in adults hospitalized with COVID-19 and evidence of lower respiratory tract infection. This trial had a total sample size of 1059 patients (538 assigned to remdesivir and 521 to placebo). The median recovery time of the remdesivir group was 11 days (95% CI [9, 12]) while that of the placebo group was 15 days (95% CI [13, 19]). The rate ratio for recovery was 1.32 (95% CI [0.47, 1.04]; *P*<.001), which was statistically significant in favor of remdesivir. The Kaplan-Meier estimates of mortality at 14 days were 7.1% with remdesivir and 11.9% with placebo, and the HR for death was (95% CI [0.47, 1.04]). Remdesivir was shown to be superior to placebo in shortening the time to recovery in adults hospitalized with COVID-19, while in terms of the HR for death, there was no significant difference between the two groups.

Given that only one treatment, remdesivir, so far has been shown to be effective by a randomized clinical trial and the pandemic of COVID-19 cannot be controlled within a short period of time, the aforementioned three clinical trials [7-9] provide extremely valuable information on the treatments of COVID-19 and the corresponding trial design and analysis. However, several important issues have been identified in the statistical analysis and implementation of the three trials. We point out the statistical issues in the three trials [7-9] and reanalyze the data from the cumulative incidence curves for the time to improvement/recovery using more appropriate approaches. Our more in-depth and comprehensive analyses yield new insights on the design and analysis for ongoing and future COVID-19 clinical trials.

## Methods

### Inflation of the type I error

The log-rank test [10] is the most commonly used method in survival analysis and clinical trial design to compare the survival benefit of two arms. Consider a randomized clinical trial with a planned sample size *N*_1_using a two-sided log-rank test. If the hypothesis test indicates no significant survival difference between the two groups under the significance level but the trial decides to continue to enroll more patients up to a larger sample size *N*_2_, this would inflate the overall type I error of the trial. Any adjustment to the sample size during the trial should be planned and evaluated in advance to maintain the overall type I error rate.

Let *Z*_1_and *Z*_2_ denote the log-rank test statistics with sample sizes *N*_1_ and *N*_2_ respectively. It holds that under the null hypothesis [11,12], and jointly follow a multivariate normal distribution,

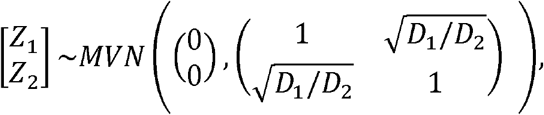

Where *D*_1_ *= dN*_1_and *D*_2_ *= dN*_2_ are the expected numbers of events with sample sizes *N*_1_ and *N*_2,_ and *d* is the proportion of patients experiencing the event. Thus, the overall type I error rate *α* _Overall_ with the significance level *α* is

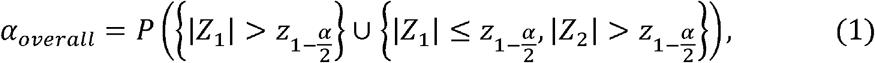

where 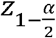 is the 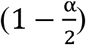 quantile of the standard normal distribution.

### Terminal (or cure) rate model

For clinical studies with survival end point, we are interested in the distribution of event time *T*. In general, patients will eventually experience the event with a long enough follow-up, although the exact event time might not be observed due to censoring. However, for some diseases with long-term survivors, it may happen that the event will never occur in a fraction of subjects, i.e., the event time for cured subjects is infinity [13-16]. Under this situation, patients can be divided into two groups: the terminal (or cure) group (the specified event would never occur) and the non-terminal group (the specified event would occur but possibly censored due to the end time of the study). Thus, the distribution of the event time *T* has a point probability mass η at ∞,

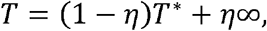

where η is the group label taking a value of 1 if the individual is in the terminal group and 0 otherwise, *γ* = *P*(η = 1) = *P*(*T =* ∞) is the terminal rate and *T*\* follows a proper and 0 otherwise, *γ* = *P*(η = 1) = *P*(*T =* ∞) is the terminal rate and *T*\* follows a proper distribution with *P*(*T*\* < ∞) =.1.For the COVID-19 trials [7, 8], the cumulative incidence curve of *T* can be expressed by

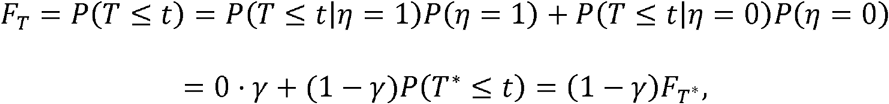

where *F*_*T*_and *F*_*T*_ * are the cumulative distribution functions of *T* and *T* *, respectively. Note that *P*(*T* < ∞) =.1 *γ* − < 1.

### Restricted mean survival time

Restricted mean survival time (RMST) [11, 17-21] is an alternative measure for the mean survival time that is not estimable due to the presence of censoring. The RMST equals to the expectation of the minimum value of event time and the specified time point τ, which can be calculated as the area under the survival curve from 0 to *τ*. It can be estimated by the area under the Kaplan-Meier survival curve, which has gained enormous popularity due to its robustness feature.

Although the HR is the most popular statistic to quantify the survival difference in randomized clinical trials, it is no longer an interpretable quantity if the proportional hazards (PH) assumption is violated [20]. By contrast, the RMST has the advantages of being nonparametric and model-free yet carrying clinically meaningful interpretations. Given the pre-specified time point τ, the estimate of the RMST difference between two groups can be interpreted as the extra survival gain on average during the time-τfollow-up period.

### Predicted trial outcome with sample size projection

Clinical trials during the epidemic of an infectious disease might fail to reach the planned sample size due to a lack of eligible patients if the outbreak can be quickly controlled [22]. However, early termination of a clinical trial would inevitably lead to loss of power and thus inconvincible findings. Based on the collected data, the bootstrap method can be used to predict what would happen if the trial had continued to reach the desired sample size. Let denote the desired sample size and *N*_*0*_(*N*_0_ < *N*) the actual number of patients enrolled. Prediction of the statistic of interest can be conducted under either conditional or unconditional schemes. The unconditional prediction draws samples (sampling with replacement from the original data with *N*_0_ observations), while the conditional prediction draws *N*_0_ − *N*_0_samples from the original *N*_0_observations and keeps the original *N*_0_ samples intact. By repeating the sampling procedure for a large number of times, one can estimate the predicted mean and the corresponding confidence interval for the statistic of interest if the trial had continued to reach the sample size of *N*.

## Results

### Lopinavir-ritonavir trial of Cao et al. [7]

In the original analysis of Cao et al. [7], the time to clinical improvement was assessed after all patients had reached day 28, while failure to reach clinical improvement or death before day 28 were considered as right-censored at day 28. In contrast to the usual survival analysis where death (or a bad event such as disease progression) is used as the event of interest, a good event (clinical improvement) was adopted as the end point in this trial. As a result, the shorter time to reach clinical improvement, the better. Cao et al. [7] concluded no benefit of using the lopinavir-ritonavir treatment beyond the standard care with an HR of 1.24 (95% CI [0.90,1.72]).

We carried out an in-depth and comprehensive investigation of the trial design in Cao et al. [7] and identified several key issues with the trial that might have hindered its success. First, the unplanned sample size increment from 160 to 199 would inflate the type I error rate. For this trial, we have *N*_1_ = 160, *N*_2_ = 199, *d* = 0.75,*D*_1_ = 160 × 0.75 = 120,*D*_2_ = 199 × 0.75 = 149.25, and based on Equation (1), *α* _*Overall*_ = 0.071 when the nominal significance level is set as *α=* 0.05. That is, the false positive rate for this trial increased as high as 7.1% in contrast to the nominal level of 5%. Any sample size alteration or re-estimation should be planned in advance in order to control the type I error rate and maintain the integrity of a trial. When the sample size reached 199, the trial was halted for enrollment because of the availability of another treatment, remdesivir. Such termination of a trial was again unplanned and immature; if there were no another agent available, would the trial continue recruitment? Interestingly, the remdesivir trial by Wang et al. (the same research group) started three days later after the lopinavir-ritonavir trial was terminated.

In terms of the primary end point, clinical improvement using two-level increment on a seven-category ordinal scale from baseline is ad-hoc due to uneven clinical differences between adjacent scales. For example, it is ambiguous whether the status of a patient changing from point 5 to point 3 is equivalent to that of changing from point 6 to point 4. In addition, live discharge from the hospital may occur from point 3 to point 2 or point 4 to point 2, which cannot be considered equivalent either. Thus, choosing 2-points improvement on the clinical outcome scale is not a precise end point, which ignores the 1-point improvement and the difference between 2-points and 3-points improvement. Instead, we recommend death as a single and clean end point for such trials, given the mortality rate was not low with severe hospitalized COVID-19 patients (19.2% in the lopinavir-ritonavir group and 25.0% in the standard care group).

The original analysis [7] treated death before day 28 as right-censored at day 28, no matter when death had occurred. This may cause ambiguity because it cannot distinguish the situations where all deaths in one group occurred earlier while those in the other group occurred later. As death is a terminal event, a terminal (or cure) rate model would be more appropriate for analysis of such data. A terminal rate model can be viewed as the counterpart of the traditional mixture cure rate model [13-16], which can be developed by slight modifications. As death is a terminal event, patients who died during the 28-day follow-up period would never reach the clinical improvement, i.e., the time to clinical improvement was infinity, denoted as ∞. Death can also be viewed as competing risk for clinical improvement.

The upper panel of Table 1 shows that neither was there any significant difference in the terminal rates between the lopinavir-ritonavir and standard care groups, nor in the HR (after excluding the terminal subjects who would eventually be absorbed in the death state) from the mixture terminal rate model. In particular, the terminal rates (including observed deaths as well as unobserved deaths that would occur after day 28 but censored at day 28) were 21.17% for the lopinavir-ritonavir group and 29.91% for the standard care group with *P*=.16, and the HR for non-terminal subjects was 1.05 (95% CI [0.78,1.42]; *P*=.74).

**Table 1.** Comparisons of estimates from the mixture terminal (or cure) model and the restricted mean time to improvement (RMTI) based on the reconstructed data^a^ from Fig. 2 in Cao et al. [7].

|  | Lopinavir–ritonavir | Standard care | Difference | *P* Value |
| --- | --- | --- | --- | --- |
| Terminal rate model<sup>b</sup> |  |  |  |  |
| Terminal rate, % | 21.17 [15.77, 28.42] | 29.91 [24.40, 36.66] | -8.74 [-21.04, 3.55] | .16 |
| Hazard ratio | 1.05 [0.78, 1.42] |  |  | .74 |
| RMTI<sup>c</sup> |  |  |  |  |
| Day 7 | 6.91 [6.79,7.00] | 6.98 [6.94,7.00] | -0.07 [-0.19, 0.05] | .26 |
| Day 14 | 12.58 [12.11, 13.04] | 13.25 [12.92,13.58] | -0.67 [-1.24, -0.11] | .02 |
| Day 28 | 17.19 [15.78, 18.60] | 18.86 [17.51, 20.21] | -1.67 [-3.62, 0.28] | .09 |

<sup>a</sup>Cumulative incidence curves were extracted and reconstructed from Fig. 2 in the Cao et al. [7] using the “digitize” package [23] in R software.
<sup>b</sup>The mixture terminal rate model was performed using the “smcure” package.
<sup>c</sup>The RMTI was estimated by calculating the area above the cumulative incidence curve using the “survRM2” package

Moreover, the crossings of the cumulative event curves for the lopinavir-ritonavir and standard care groups at days 10 and 16 in Fig. 2 [7] imply possible violation of the PH assumption. When the PH assumption is not satisfied, the HR from a Cox model [24] is not clinically meaningful. As an alternative, the area above the curve in Fig. 2 [7] or the area under the inverted curve as shown in our Fig. 1, referred to as the restricted mean time to improvement (RMTI), can be used to quantify treatment effect which requires no assumption such as PH [11, 17-21]. As a model-free quantity, the RMTI up to 28 days can be interpreted as the average time to reach improvement in 28 days, for which the shorter the better. The 28-day RMTI difference between the two groups was −1.67 days (95% CI [-3.62, 0.28]; *P*=.09) in favor of lopinavir-ritonavir but not statistically significant. The 7-day and 14-day RMTIs are also presented in the lower panel of Table 1, where the 14-day RMTI showed some promising results for lopinavir-ritonavir yet further confirmation is needed.

**Figure 1.**
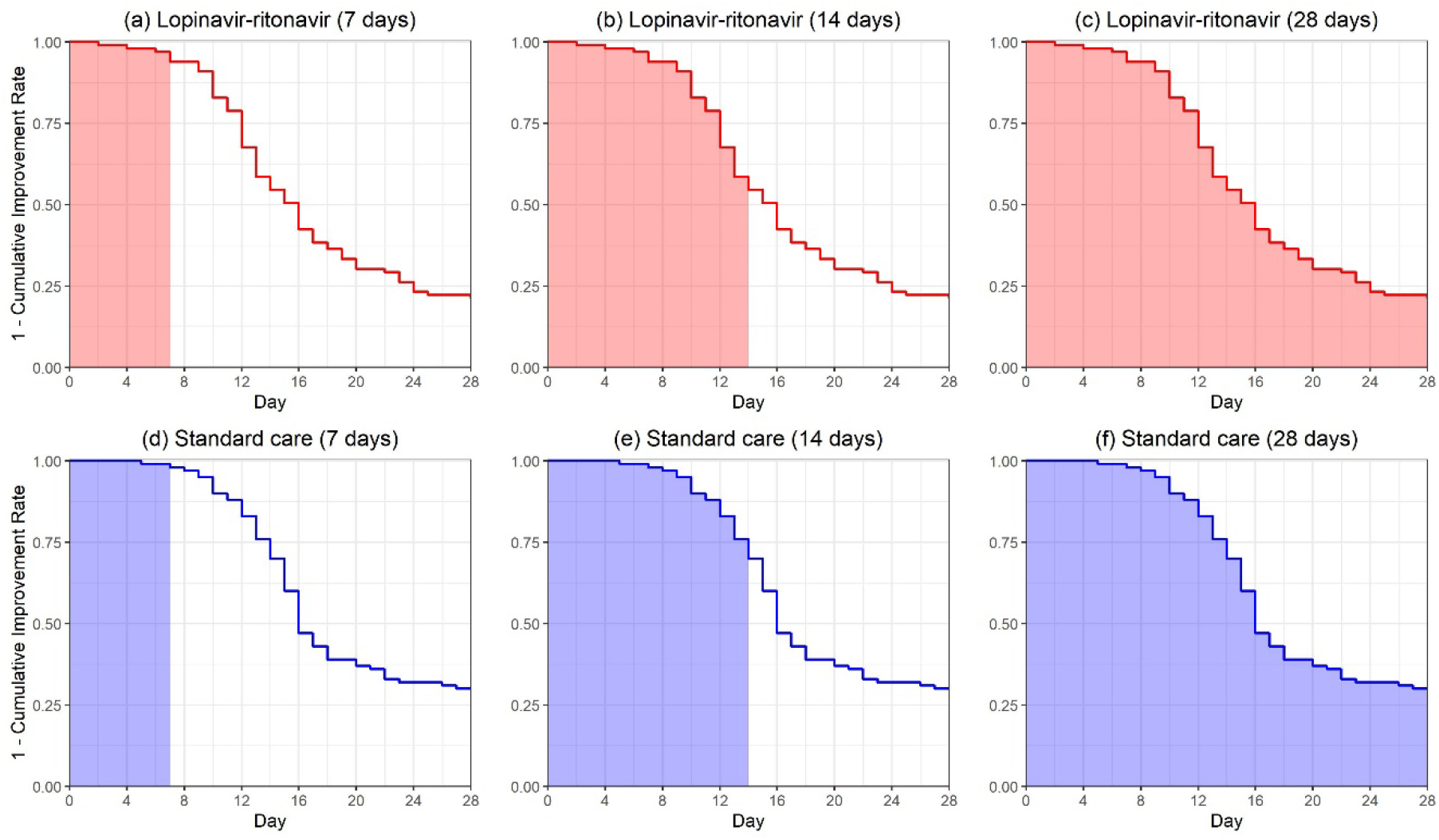
The restricted mean time to improvement (RMTI) corresponding to the area under the curves for the lopinavir-ritonavir group and the standard care group evaluated at days 7, 14, 28 in Cao et al. [7].

**Figure 2.**
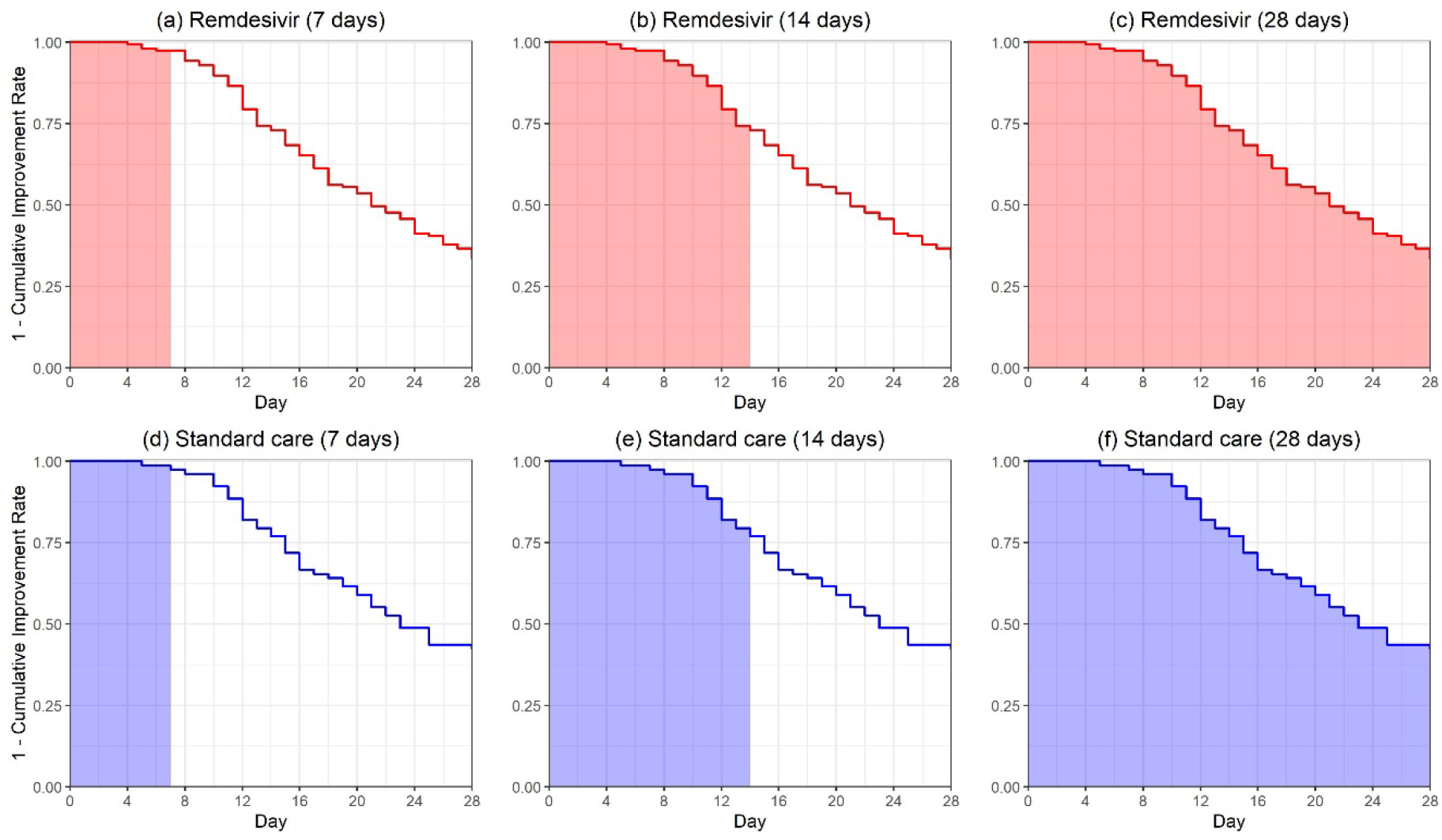
The restricted mean time to improvement (RMTI) corresponding to the area under the curves for the remdesivir group and the placebo group evaluated at days 7, 14, 28 in Wang et al. [8].

Tables 2 and 3 show the numbers on mortality and clinical improvement by day 28 across the two treatment groups respectively. We carry out chi-squared tests (or Fisher’s exact tests if some of the cell counts are smaller than 5) to examine any association between the outcomes and treatments. For Table 2 with 2×3 cells, there is no association with *P*=.53, and if combining deaths in both earlier and later stages, this leads to 2×2 cells with *P*=.32 and odds ratio 0.71 (95% CI [0.36, 1.40]). Patients treated with lopinavir-ritonavir had times odds to die by day 28 in comparison to those with the standard care group. For Table 3 with 2×4 cells, there is no association with *P*=.11, and if combining all clinical improvement cases, this leads to 2×2 cells with *P*=.53 and odds ratio 1.24 (95% CI [0.64, 2.40]). Patients treated with lopinavir-ritonavir had 1.24 times odds to achieve clinical improvement by day 28 in comparison to those with the standard care group. However, none of the results are statistically significant.

**Table 2.** Counts of deaths for the earlier stage (≤12 days after onset of symptoms) and later stage (>12 days after onset of symptoms) and survivors.

|  | Deaths |  | Survivors |
| --- | --- | --- | --- |
| Treatment | Earlier | Later |  |
| Lopinavir-ritonavir | 8 | 11 | 80 |
| Standard care | 13 | 12 | 75 |

**Table 3.** Counts of clinical improvement cases in days 1 to 7, 8 to 14, 15 to 28 and non-improvement cases.

|  | Clinical Improvement |  |  | No improvement |
| --- | --- | --- | --- | --- |
| Treatment | Days 1 to 7 | Days 8 to 14 | Days 15 to 28 |  |
| Lopinavir-ritonavir | 6 | 39 | 33 | 22 |
| Standard care | 2 | 28 | 40 | 30 |

### Remdesivir trial of Wang et al. [8]

Wang et al. [8] reported a randomized, double-blind, placebo-controlled trial for remdesivir with severe COVID-19 patients. Based on an adjusted six-point ordinal scale of clinical status, the primary end point was the time to clinical improvement, defined as 2-level decline from randomization (similar to that in Cao et al. [7], in fact the two trials were conducted by the same group of investigators), for which the shorter the better. Patients were permitted concomitant use of lopinavir-ritonavir, interferons, and corticosteroids. The HR between the remdesivir and placebo groups was 1.23 [95% CI 0.87-1.75], indicating no significant difference. Overall, 237 eligible patients were enrolled, with 158 patients assigned to the remdesivir group and 78 patients to the placebo group under the intent-to-treat (ITT) scheme. The trial was stopped early and thus failed to reach the designated sample size 453 due to a lack of eligible patients.

Similar to the trial by Cao et al. [7], deaths before day 28 were treated as right-censored observations at day 28 regardless the actual occurrence time of deaths in Wang et al. [8]. Moreover, a clinical improvement might not be observed due to death, i.e., death is a terminal event, and thus the terminal (or cure) rate model introduced earlier should be recommended for the survival analysis rather than the standard Cox model.

The upper panel of Table 4 indicates no significant difference in the terminal rates between the remdesivir and placebo groups. In particular, the terminal rates were 31.49% for the remdesivir group and 40.71% for the placebo group with *P*=.19. With the terminal subjects excluded, the HR from the mixture terminal rate model was 0.92 (95% CI, [0.63, 1.35]; *P*=.67), which also showed no significant difference between the two groups.

**Table 4.** Comparisons of the estimates from the mixture terminal (or cure) rate model and the restricted mean time to improvement (RMTI) based on the reconstructed data from Fig. 2 in Wang et al. [8].

|  | Remdesivir | Placebo | Difference | *P* Value |
| --- | --- | --- | --- | --- |
| Terminal rate model |  |  |  |  |
| Terminal rate, % | 0.31 [0.27, 0.37] | 0.41 [0.32, 0.51] | -9.22 [-22.9, 4.45] | .19 |
| Hazard ratio | 0.92 [0.63, 1.35] |  |  | .67 |
| RMTI |  |  |  |  |
| Day 7 | 6.95 [6.90, 7.00] | 6.97 [6.92, 7.00] | -0.03 [-0.10, 0.05] | .49 |
| Day 14 | 13.09 [12.78, 13.40] | 13.29 [12.92, 13.67] | -0.20 [-0.69, 0.29] | .42 |
| Day 28 | 20.42 [19.26, 21.57] | 21.31 [19.73, 22.88] | -0.89 [-2.84, 1.06] | .37 |

Due to the competing risk from death, the end point might not be observed, and thus the standard hazard concept is ambiguous and the HR does not have a meaningful interpretation anymore [25]. In Fig. 2 [8], the curve for the cumulative improvement event of remdesivir is uniformly higher than that of the control, indicating patients with remdesivir reached improvement faster than those in the control group. The area above the cumulative incidence curve or equivalently the area under the survival curve up to 28 days in our Fig. 2 would be a reasonable quantity for evaluating the treatment efficacy.

Using the reconstructed data from Fig. 2 [8], the restricted mean time to improvement (RMTI) evaluated at day 28 was 20.42 days (95% CI [19.26, 21.57]) for the remdesivir group and 21.31 days (95% CI [19.73, 22.88]) for the placebo group. As shown in the lower panel of Table 4, the difference in RMTIs was −0.89 day (95% CI [-2.84, 1.06]), numerically favoring remdesivir but not statistically significant. It can be interpreted that patients treated by remdesivir on average enjoyed extra 0.89 day of improvement during the 28-day follow-up compared with those in the placebo group. The 7-day and 14-day RMTIs are also presented in the lower panel of Table 4, while neither showed statistically significant results.

The trial was terminated without reaching the originally planned sample size 453 due to a lack of eligible patients. With only 236 patients in the ITT analysis, the estimated HR was 1.23 (95% CI [0.87, 1.75]), numerically favoring remdesivir, which however might not be reliable due to the underpowered study. Using the bootstrap method, we can predict what would happen if the trial had continued to reach the full sample size or double the planned sample size. Table 5 shows both the unconditional and conditional predictions of the HR, similar to sample size re-estimation using conditional power [26] in a two-stage design. If the trial could have reached the designated sample size, the HR from the conditional prediction shows significant treatment effect of remdesivir with *P*=.02, and if the trial could enroll twice of the target sample size, both conditional and unconditional approaches result in significant differences under the 5% significance level. Thus, a larger sample size may be needed to show the significant difference between remdesivir and placebo.

**Table 5.** Predicted hazard ratios (with 95% confidence intervals) and *P* values at the actual, target, and double of the target sample sizes using 50000 bootstrap samples based on the reconstructed data from Fig. 2 in Wang et al. [8].

| □ Sample size | Sample size in each arm |  | Unconditional Prediction |  | Conditional Prediction |  |
| --- | --- | --- | --- | --- | --- | --- |
|  | Remdesivir | Placebo | HR [95% CI] | *P* value | HR [95% CI] | *P* value |
| Actual | 158 | 78 | 1.23 [0.87, 1.75] | .24 | □ | □ |
| Target | 302 | 151 | 1.24 [0.96, 1.60] | .10 | 1.24 [1.03, 1.48] | .02 |
| Target×2 | 604 | 302 | 1.24 [1.03, 1.48] | .02 | 1.24 [1.06, 1.44] | .01 |

### Remdesivir trial of Beigel et al. [9]

Beigel et al. [9] presented a preliminary report of NCT04280705 trial which is a randomized, double-blind, placebo-controlled trial of intravenous remdesivir in adults hospitalized with COVID-19 with evidence of lower respiratory tract involvement. This trial enrolled 1059 patients (538 assigned to remdesivir and 521 to placebo). The primary end point of the original analysis was the recovery time, defined by either discharge from the hospital or hospitalization for infection-control purposes only. The median recovery time of the remdesivir group was 11 days (95% CI [9, 12]) and that of the placebo group was 15 days (95% CI [13, 19]). The rate ratio of recovery for remdesivir vs. placebo was 1.32 (95% CI [1.12, 1.55]; *P*<.001), which demonstrated the superiority of remdesivir. In terms of the HR for death, there was no significant difference between the remdesivir and placebo groups with an HR of 0.70 (95% CI [0.47, 1.04]).

The remdesivir trial of Beigel et al. [9] is essential to evaluate the efficacy of remdesivir, as it has a large sample size of 1059 patients under a well-designed randomized controlled trial scheme. In terms of the data analysis, Beigel et al. [9] only reported the median recovery time without *P* value. From Fig. 2A [9], the Kaplan-Meier curves of cumulative recoveries are initially intertwined and then diverge, so other percentiles of the time to recovery would provide more information on the efficacy of remdesivir. Meanwhile, a global and robust measurement, the restricted mean time to recovery (RMTR), can help to quantify the treatment efficacy in a more comprehensive way [11,17-21].

The upper panel of Table 6 presents the RMTRs up to day 30 for both remdesivir and placebo groups. The RMTRs were 14.5 days and 17.2 days for remdesivir and placebo respectively, indicating that patients with remdesivir on average enjoyed 2.7-days gain of recovery with 30-days follow-up. The difference in RMTRs was statistically significant with *P*<.001, demonstrating the superiority of remdesivir. This is consistent with the original analysis in terms of the rate ratio of recovery [9]. Meanwhile in the bottom penal of Table 6, more percentiles of the time to recovery were reported with *P* values. The early difference for remdesivir vs. placebo in the recovery time at the 25th percentile was −1 (95% CI [−3, 0]; *P*=.65), which was not statistically significant. However, the differences manifested to be statistically significant later; for example, the 30th to 60th percentiles of the recovery time in the remdesivir group were all significantly shorter than those in the placebo group. It is reasonable for the treatment to take effect after a certain length of follow-up.

**Table 6.** The restricted mean time to recovery (RMTR) and percentiles of the time to recovery based on the reconstructed data from Fig. 2A of Beigel et al. [9].

| Statistical measure | Remdesivir | Placebo | Difference<br>[95% CI] | $P$ Value |
| --- | --- | --- | --- | --- |
| RMTR (up to day 30) | 14.5<br>[13.6, 15.5] | 17.2<br>[16.1, 18.2] | -2.7<br>[-4.0, -1.2] | <.001 |
| Percentiles of the time to recovery [95% CI] |  |  |  |  |
| 25th | 5 [4, 5] | 6 [6, 7] | -1 [-3, 0] | .649 |
| 30th | 6 [5, 6] | 8 [7, 9] | -2 [-4, -1] | .002 |
| 40th | 8 [7, 9] | 11 [9, 13] | -3 [-5, -1] | .007 |
| 50th (median) | 11 [9, 12] | 15 [13, 19] | -4 [-9, -2] | .010 |
| 60th | 15 [13, 19] | 22 [20, 27] | -7 [-12, -3] | .004 |

## Discussion

When designing and conducting a clinical trial for new treatment, particularly for the COVID-19 pandemic without knowing much about the clinical outcomes, many things can go wrong if the design is not well thought through, the trial is not carefully conducted following the protocol, or the analysis is not properly carried out. Critical issues with such trials include, but not limited to, the end point selection, the type I error rate control, double blinding or open label, early termination of a trial, the validity of the PH assumption in a Cox model, assumptions for statistical tests and models, etc. In contrast to searching for a needle in a haystack, the trial design should be more targeted, focused, and tailored for specific needs of COVID-19 patients and particular disease characteristics and severities [27].

Given the emergency and fast spread of the coronavirus around the world, it is crucial to design the right clinical trial and accelerate the development of new treatment. With the high speed of enrollment and urgency of the trial outcome, it appears to be difficult to carry out any adaptation during the trial conduct. The trial outcomes unfold so fast that any adaptation may not be able to catch up with the speed of recruitment.

As a summary, our recommendations for COVID-19 trials are listed as follows:

1. Adopt death as a single end point for severe hospitalized COVID-19 patients; or live discharge from the hospital for moderately severe COVID-19 patients.
2. Conduct the gold standard trial scheme: a randomized, double-blind, controlled trial with equal randomization, 1:2 or 1:3 allocation ratio for control vs. treatment.
3. With multiple agents tested in one trial, allow the trial to drop certain treatment due to futility or toxicity.
4. Adopt the restricted mean survival time as the metric to quantify the treatment effect when the proportional hazards assumption is not satisfied; otherwise standard approaches using the hazard ratios and log-rank tests should be used.
5. Control the type I error rate; any sample size alternation during the trial must be planned and evaluated in advance with a strict control of the false positive rate.
6. Intent-to-treat analysis (or its modified version) is recommended for the final analysis.

Although adaptive design has gained much popularity and is playing an increasingly important role in oncology trials, the advantages of adaptive design may be mitigated to large extent under such a fast patient enrollment, because the impact of any adaptation may be too slow to manifest before the trial is completed. As a result, our recommendations follow the gold standard scheme of conventional trial design without much adaptation ingredient, which may help investigators to discriminate different treatments and identify the effective ones in an efficient way.

## Data Availability

The authors confirm that the data supporting the findings of this study are available within the article.

## Notes

### Competing Interest Statement

The authors have declared no competing interest.

### Funding Statement

No funding to disclose.

### Author Declarations

IRB exemption category 4 - Existing data

